# Prevalence of anxiety and depressive symptoms among physicians during the COVID-19 pandemic in Bangladesh: a cross-sectional study

**DOI:** 10.1101/2020.12.08.20245829

**Authors:** M.Tasdik Hasan, Sahadat Hossain, Farhana Safa, Afifa Anjum, Abid Hasan Khan, Kamrun Nahar Koly, Syeda Fatema Alam, Md. Abdur Rafi, Vivek Podder, Tonima Islam Trisa, Rhedeya Nury Nodi, Dewan Tasnia Azad, Fatema Ashraf, S.M Quamrul Akther, Helal Uddin Ahmed, Simon Rosenbaum, Graham Thornicroft

## Abstract

**Objectives:** In addition to risking their physical well-being, frontline physicians are enduring significant emotional burden both at work and home during the COVID-19 pandemic. This study aims to investigate the levels of anxiety and depressive symptoms and to identify associated factors among Bangladeshi physicians during the COVID-19 outbreak.

**Methods and design:** A cross-sectional study using an online survey was conducted between April 21 and May 10, 2020. Outcomes assessed included demographic questions, COVID-19 related questions, and the Hospital Anxiety and Depression Scale (HADS).

**Results:** The survey was completed by 412 Bangladeshi physicians. The findings revealed that, in terms of standardized HADS cut-off points, the prevalence of anxiety and depressive symptoms among physicians was 67.72% and 48.5% respectively. Risk factors for higher rates of anxiety or depressive symptoms were: being female, physicians who had experienced COVID-19 like symptoms during the pandemic, those who had not received incentives, those who used self-funded PPE, not received adequate training, lacking perceived self-efficacy to manage COVID -19 positive patients, greater perceived stress of being infected, fear of getting assaulted/humiliated, being more connected with social media, having lower income levels to support the family, feeling more agitated, less than 2 hours of leisure activity per day and short sleep duration. All these factors were found to be positively associated with anxiety and depression in unadjusted and adjusted statistical models.

**Conclusions:** This study identifies a real concern about the prevalence of anxiety and depressive symptoms among Bangladeshi physicians and identifies several associated factors during the COVID-19 pandemic. Given the vulnerability of the physicians in this extraordinary period whilst they are putting their own lives at risk to help people infected by COVID-19, health authorities should address the psychological needs of medical staff and formulate effective strategies to support vital frontline health workers.

**STHRENGHTS & LIMITATIONS OF THE STUDY:**

- This study reports a novel and concerning findings on the prevalence of anxiety and depression symptoms with identification of several important associated factors among Bangladeshi physicians during the COVID-19 pandemic.
- The cross-sectional nature of the study design could not establish causal relationship between the dependent and independent variables.
- This study was carried out by conducting a web-based survey, which might generate sampling bias by excluding the physicians who do not have access to internet or inactive in social medias, and thus limit the generalizability of the findings.

## INTRODUCTION

The unprecedented and unpredictable nature of the COVID-19 pandemic has triggered a focus on the psychological and mental health problems of the health care staff involved. ^1,2^ Physicians being key frontline workers are among the most affected of health care staff professions. A cross-sectional study based on an investigation involving 34 hospitals in China reported that frontline healthcare workers often experienced depressive symptoms, anxiety, sleep disturbances, and distress whilst managing patients with COVID-19.^3^ The situation is similar in many countries and particularly challenging in low-resource settings.^2^

The first case of COVID-19 in Bangladesh was announced on 08 March 2020 and the first death was documented on 18 March 2020.^4^ Over time, the number increased to 2,97,083 as reported on 24^th^ August 2020 with a death toll of 3,983 patients^5^ whilst the total number of affected physicians was 7843 with a tragic loss of 88 doctors.^6^ Ing and colleagues (2020) attempted to enumerate the total number of physician deaths around the world until mid-April 2020, with a reported 278 deaths. However, this number has been shown to be increasing since then.^7^ In Bangladesh, a severe dearth in resources and support have been observed since the initial phase of the pandemic for front line physicians.^8^ Hassan et al., (2020) expressed the concerns of Bangladeshi physicians in terms of infecting their own families, considering the tradition of congested, multifamily accommodation with limited quarantine opportunities. They also described, in addition to risking their own physical well-being, that frontline physicians are enduring significant emotional burdens both at work and at home.^9^

Therefore, it is essential to identify and characterize the mental health difficulties experiencing by the Bangladeshi physicians during the pandemic in such a challenging setting. There is no published evidence on mental health issues among Bangladeshi physicians related to COVID-19. This is especially pertinent with the uncertainty surrounding an outbreak of such unparalleled magnitude in such a low resource country. Therefore, this study aims to investigate the levels of anxiety and depressive symptoms and to identify associated factors among Bangladeshi registered physicians during the COVID-19 outbreak.

## MATERIALS AND METHODS

### Study design and participants

A cross sectional study was conducted among Bangladeshi registered physicians from April 21 to May 10, 2020, when the COVID-19 pandemic and the enforced lockdown was in its initial phase in the country. To be eligible, the respondents had to be adults (>18 years), Bangladeshi registered physicians, able to read and understand English, and to be living in Bangladesh at the time of the COVID19 outbreak. The Sample size was calculated from prevalence estimate using following formula: 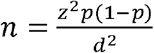, where, where n = number of sample; z = 1.96 for 95% confidence level (CI), p = “best guess” for prevalence and d = precision of the prevalence estimate. However, a recent study by Banna et al., (2020) reported that the prevalence of anxiety symptoms and depressive symptoms among general population was 33.7% and 57.9% during COVID-19 pandemic in Bangladesh.^10^ We assumed that the psychological difficulties might be 50% among the physicians of Bangladesh and so the calculated sample size was 384 participants. Assuming 15% non-response rate we calculated the sample size as 442.

We used convenience sampling method to identify & recruit appropriate participants. Considering the risky data collection inside the hospital setting amid the pandemic, an online survey was posted on closed social media (Facebook) groups of registered physicians of Bangladesh and open request was placed by the team of investigators to complete the survey. Also, five volunteers (medical doctors) from different medical institutions were employed to circulate details of the survey among their professional networks in addition to regular posting in social media groups. They were instructed to be inclusive, open and to circulate details of the survey periodically for maximum reach. Email addresses of the participants were collected upon proper clarification and informed, written consent was obtained.

The study was conducted following the Checklist for Reporting Results of Internet ESurveys (CHERRIES) guideline.^11^ The authors ensured that all procedures contributing to this work comply with the ethical standards of the relevant national and institutional committees on human experimentation and with the Helsinki Declaration of 1975, as revised in 2008. The study was approved by the Ethical Review Committee, Shaheed Suhrawardy Medical College, Dhaka, Bangladesh (ShSMC/Ethical/2020/12).

### Data collection tool

Data were collected using a structured online questionnaire created in Google form (in English). The questionnaire had 3 parts: (i) demographic questions, (ii) COVID19-related questions, (iii) Hospital Anxiety and Depression Scale (HADS; higher scores on the subscales indicate higher levels of depression and anxiety symptoms)).^12^

### Statistical analysis

All statistical analyses were carried out using SPSS (version 25.0). Frequency distribution with percentage was used to present categorical variables while mean with standard deviation (SD) was used to present continuous variables. Chi-square (χ^2^) test was used to determine any difference between groups. Both bivariate and multiple logistic regression models were used to find out the predictors of anxiety and depressive symptoms among Bangladeshi physicians during COVID-19 pandemic. Statistical significance level was set at p-value <0.05 and 95% confidence interval (CI).

## RESULTS

A total of 412 participants (response rate 93.21%) took part in the study and completed the survey. Most physicians were female (55.8%), the majority were aged between 25 to 34 years (76.2%), and most were unmarried (55.6%). More than half of the study participants (52.9%) reported having an income of 40,000 BDT (365 GBP) per month or less (see Table 1).

**Table 1:** Sociodemographic characteristics of the study participants.

|  | Variables | Frequency | Percentage |
| --- | --- | --- | --- |
| **Gender** |  |  |  |
|  | Male | 182 | 44.2 |
|  | Female | 230 | 55.8 |
| **Age** |  |  |  |
|  | ≤ 24 yrs | 67 | 16.3 |
|  | 25-34 yrs | 314 | 76.2 |
|  | ≥ 35 yrs | 31 | 7.5 |
| **Marital status** |  |  |  |
|  | Married | 173 | 42.0 |
|  | Unmarried | 229 | 55.6 |
|  | Divorced/separated | 10 | 2.4 |
| **Income (n=268)** |  |  |  |
|  | ≤ 20,000 | 125 | 30.3 |
|  | 21,000-40,000 | 93 | 22.6 |
|  | 41,000-70,000 | 31 | 7.5 |
|  | ≥ 70,000 | 19 | 4.6 |

The study showed that females (75.2%) suffered more from anxiety than males (58.2%) which was statistically significant ((p<0.001). Similarly, depression was more prevalent among females (53.9% vs. 41.8% male, p=0.014).Addition to that, Respondents (77.8%) experiencing COVID-19 symptoms were suffering from anxiety than those not experiencing symptoms of COVID-19 (65.7%) that was statistically significant (p=0.017). Anxiety was more prevalent among respondents those feel that they were not provided with training about COVID-19 (71.9%) than respondents those feel that they were provided enough training about COVID-19 (61.3%) and it was found statistically significant (p=0.025).Moreover, respondents those were not ready to deal a COVID-19 positive patient suffered more from anxiety (74.8% vs. 60.4% respondents ready to deal a COVID-19 positive patient, p=0.002). Furthermore, depression was also more prevalent among respondents not ready to deal a COVID-19 positive patient (56.7% vs. 40.1% respondents ready to deal a COVID-19 positive patient, p=0.001). Anxiety was more common among respondents severely tensed about being infected by COVID-19 (81.60% vs. 31.70% respondents not tensed at all or minimally tensed, p<0.01). In the same way, depression was also seen to be common among respondents severely tensed about being infected by COVID-19 (57.10% vs. 26.80% respondents not tensed at all or minimally tensed, p<0.01). However, variables such as receiving treatment for other diseases, and having knowledge about someone tested positive for COVID-19 had no statistically significant relationship with anxiety or depression (see Table 2).

**Table 2:**
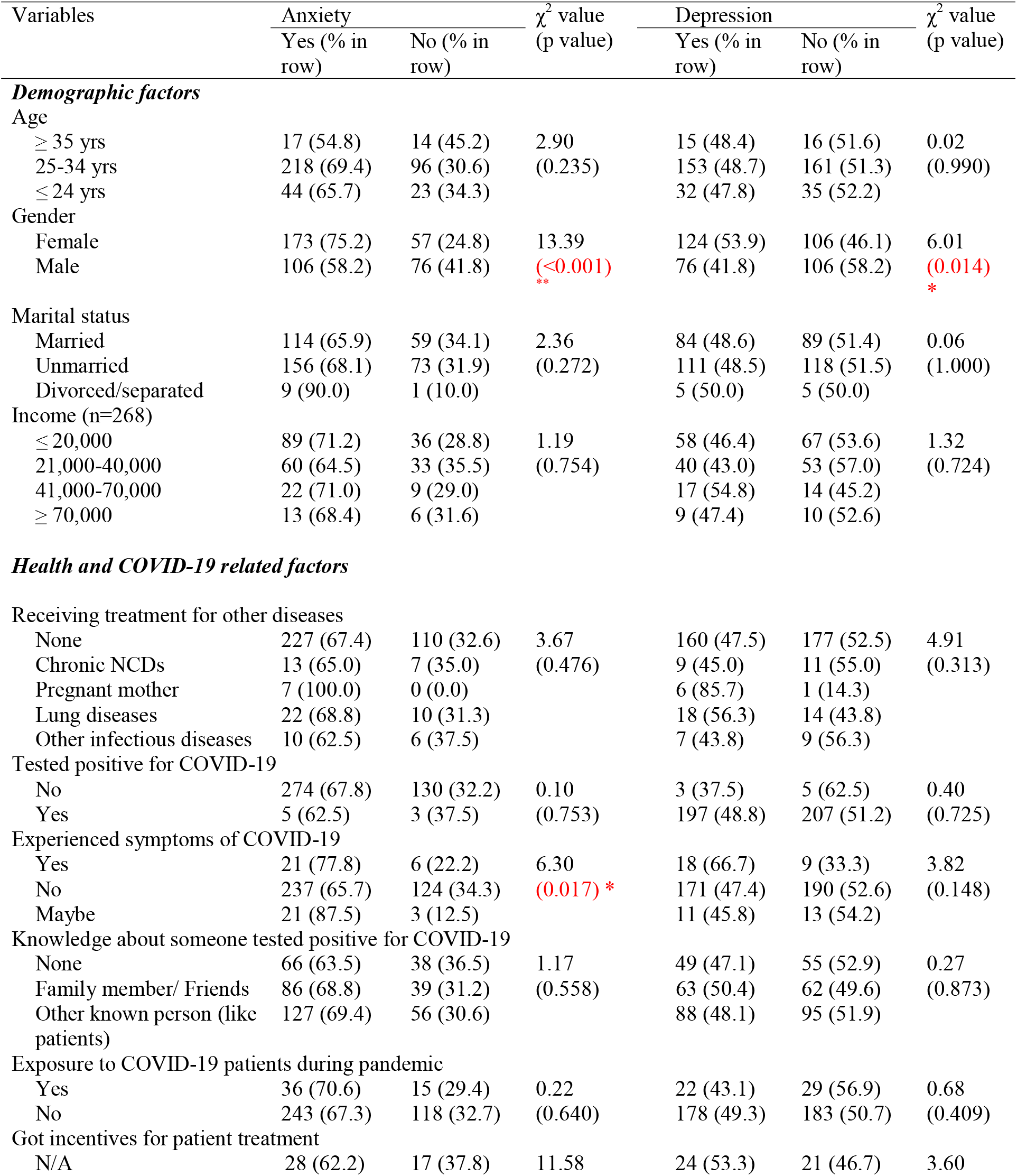
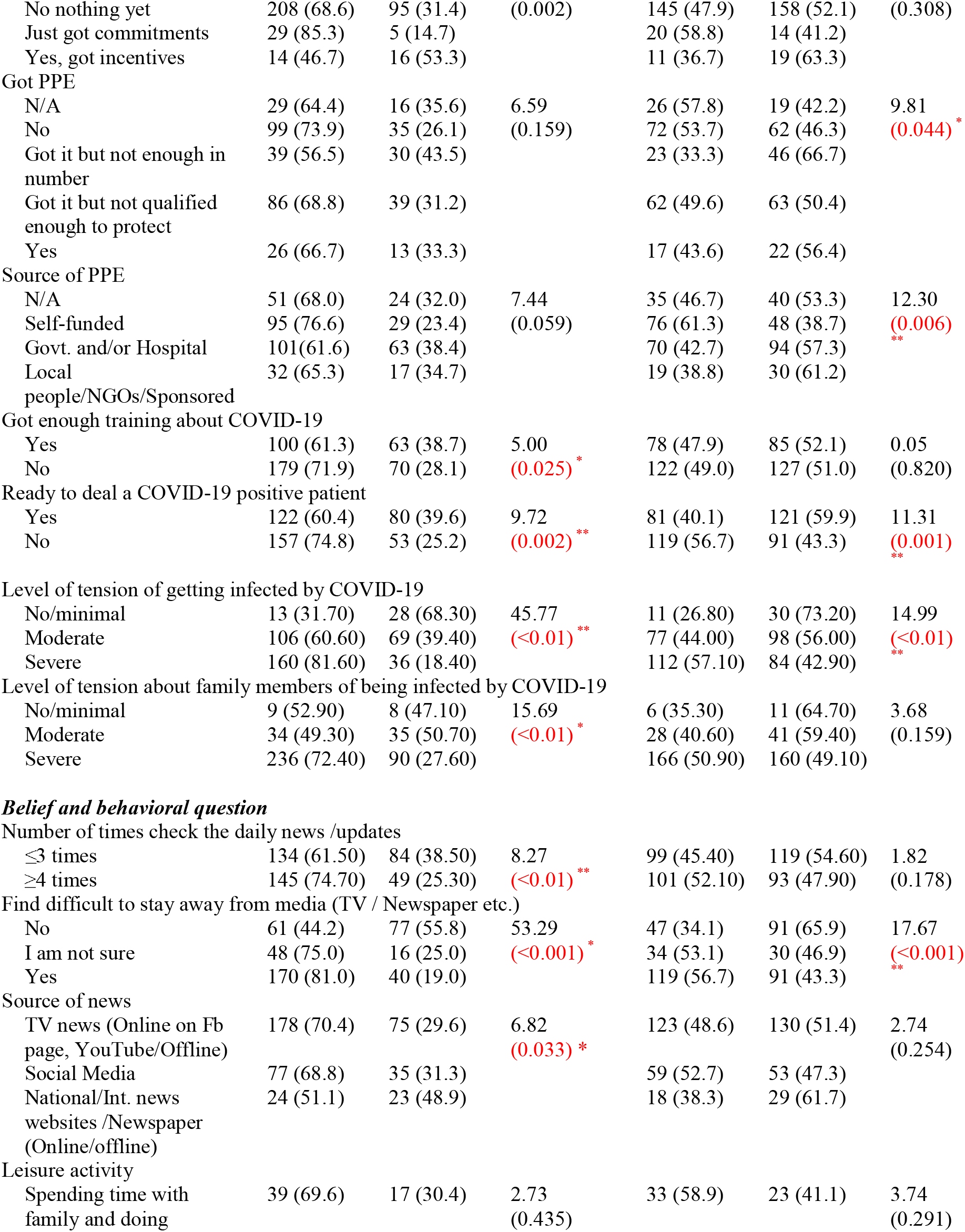
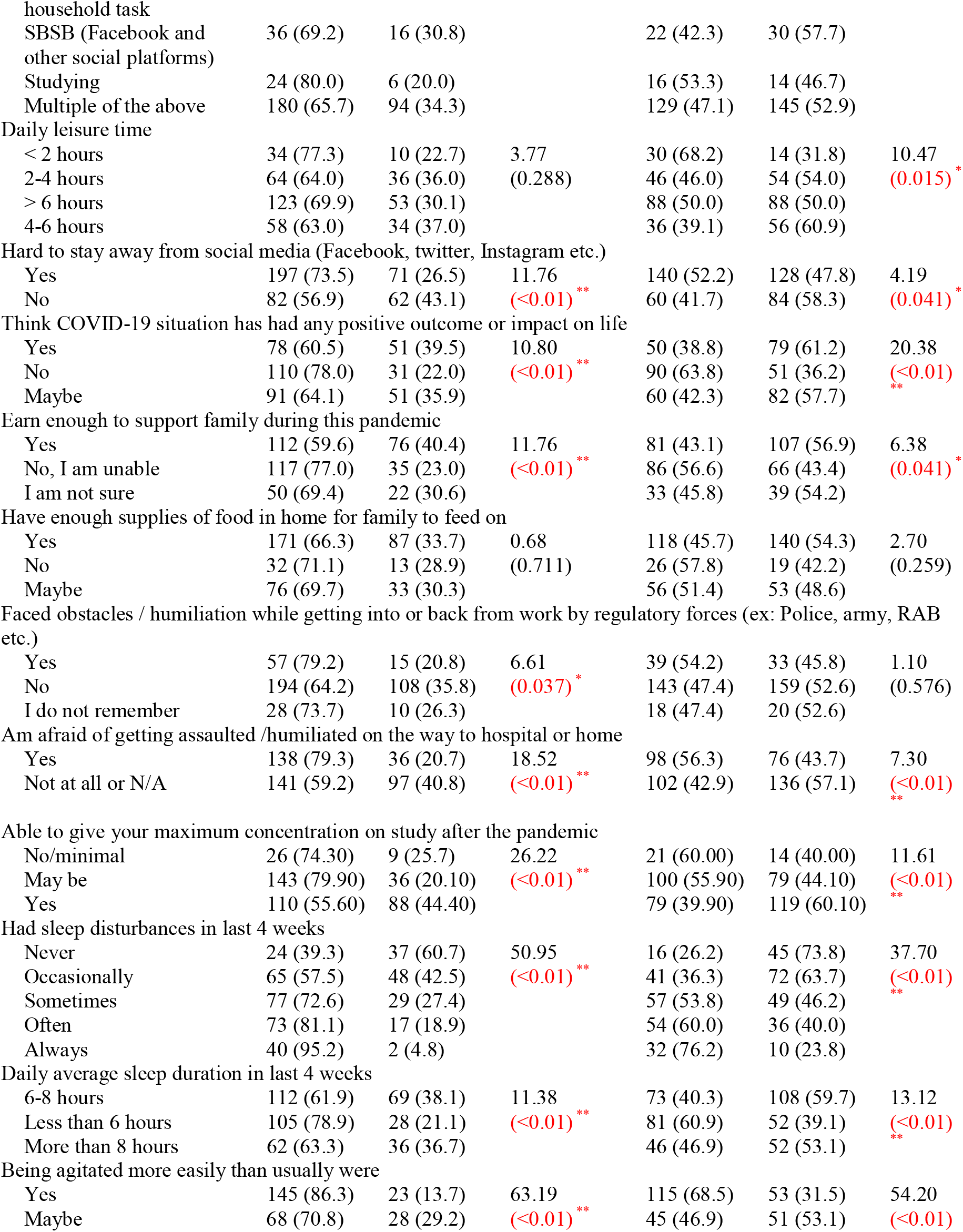
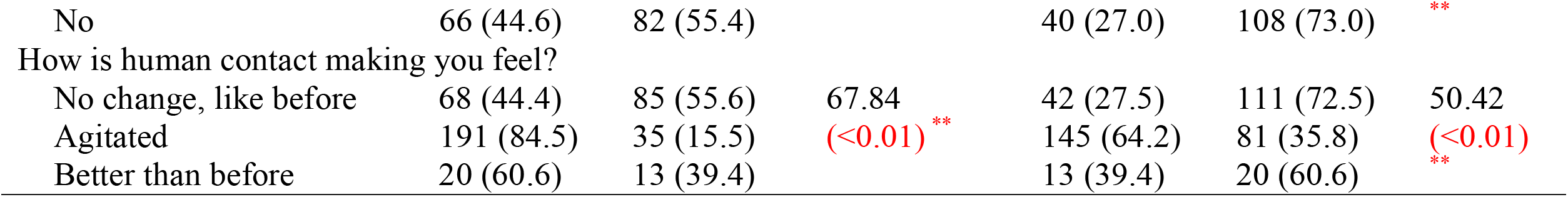
Factors associated with anxiety and depression among physicians in Bangladesh.

The study also showed that respondents checking news updates more than four times a day, having hard time staying away from media (e.g. TV, newspaper etc.), having less than 2 hours of leisure, being unable to earn enough to support the family, facing any obstacles or humiliation by regulatory forces (e.g. Police, Rapid Action Battalion etc.) on the way to work from home and vice versa, being agitated more easily than usual, being agitated with human contact had higher occurrence of either anxiety or depression or both in some cases (see Table 2).

Regression analysis showed that, females were about 2.5 times more likely to be in anxiety than males (p<0.01). Addition to that, respondents experiencing COVID-19 had higher odds to suffer from depression than respondents not experiencing COVID-19 symptoms (OR=1.63; 95% CI: 1.10-2.42).Furthermore, it was seen that respondents spending less than 2hours a day for leisure activity were about 4 times more likely to suffer from depression than respondents spending 4 to 6 hours for leisure activity (p<0.01).Moreover, respondents those felt agitated by human contact had higher likelihood for anxiety (AOR=2.68; 95% CI:1.23-5.81) and depression (AOR=2.78; 95% CI:1.50-5.16) than those not feeling any changes by human contact. Similarly, respondents being unable to give maximum concentration on job after this pandemic, having fear of getting assaulted or humiliated on the way to work or home, having no positive outcome or impact on life through this pandemic, having hard time to stay away from social media (e.g. Facebook, Twitter, Instagram etc.), sleeping less than 6 hours had significant association with higher odds of both anxiety and depression (Table 3).

**Table 3:**
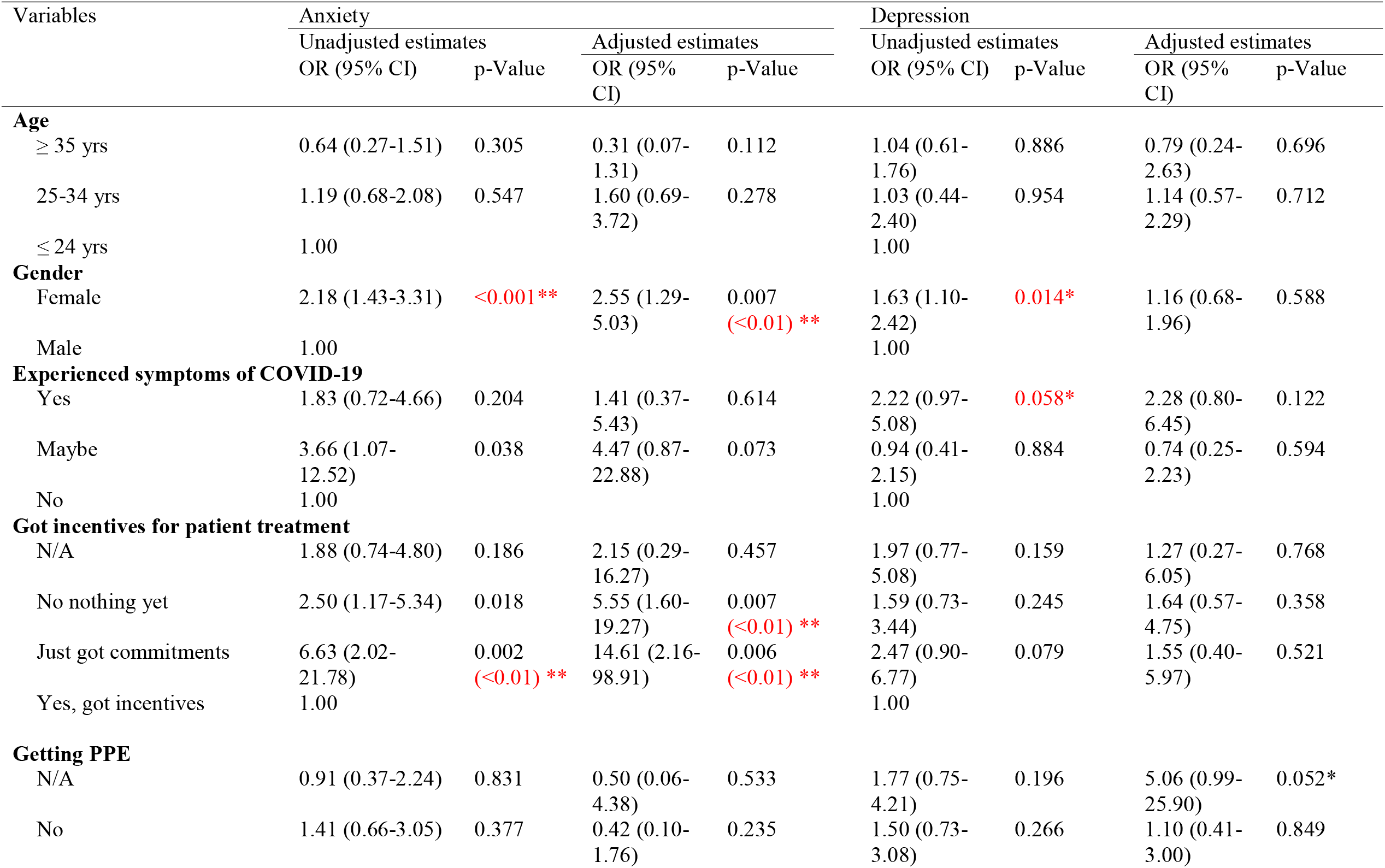
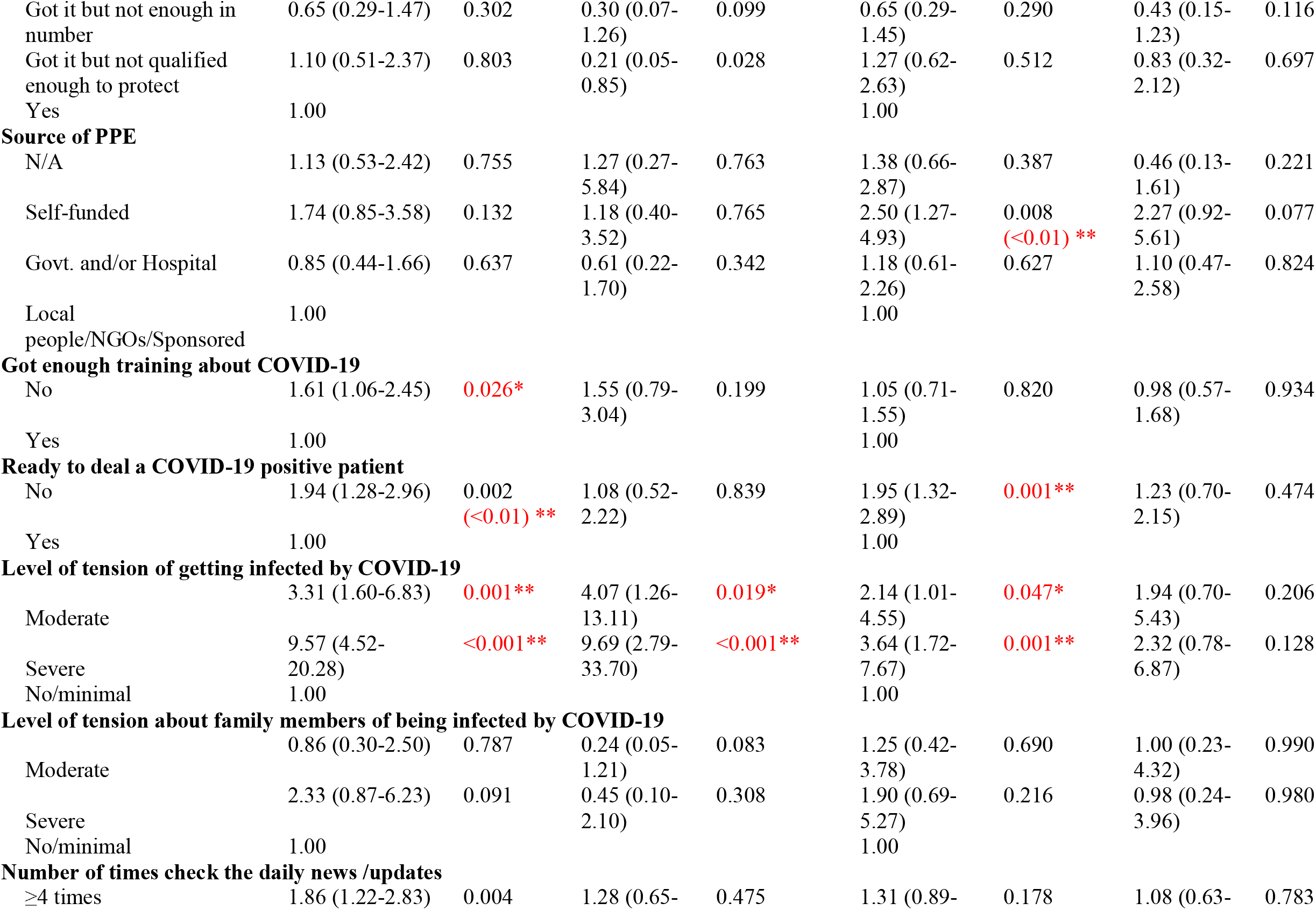
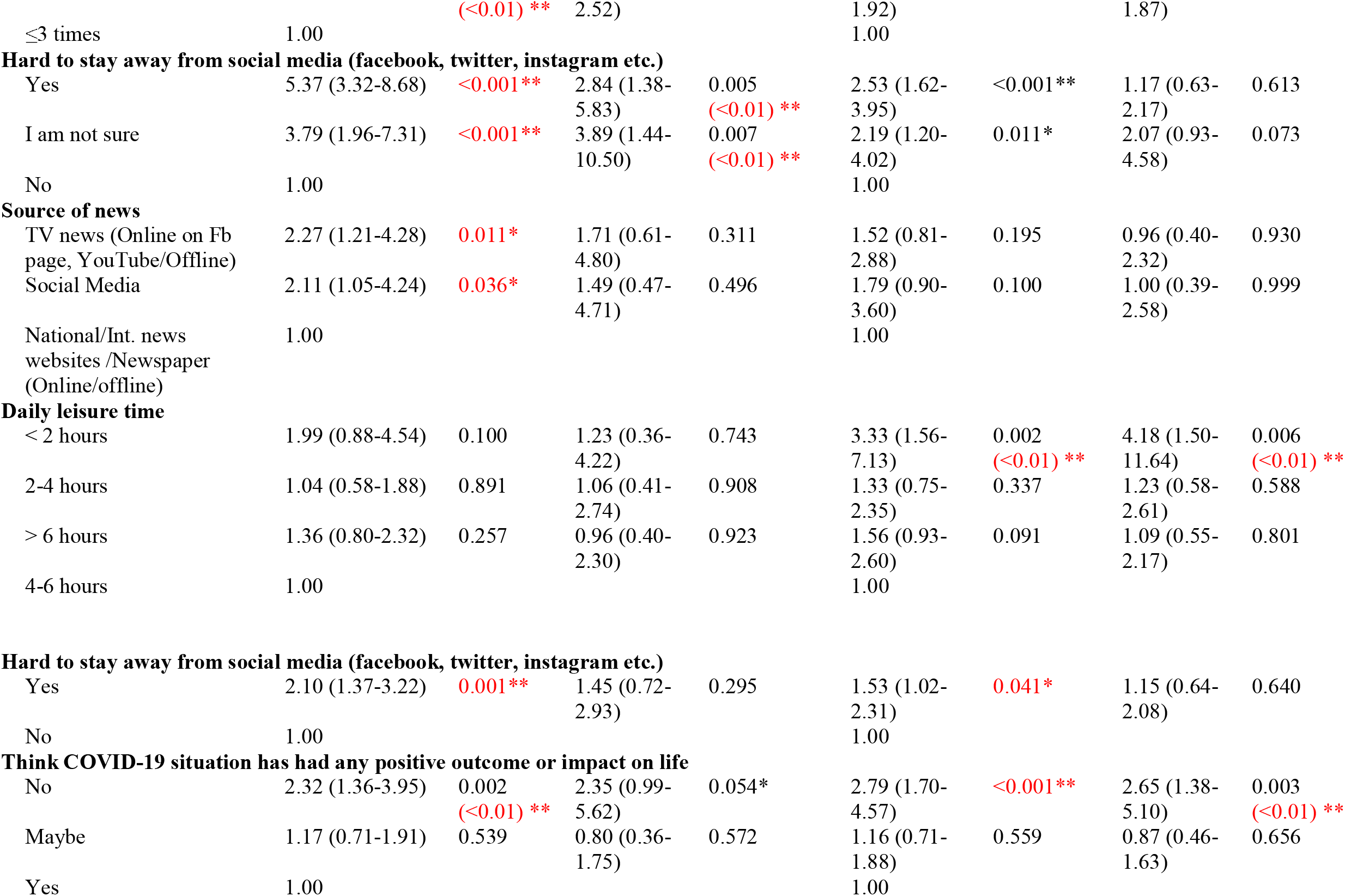
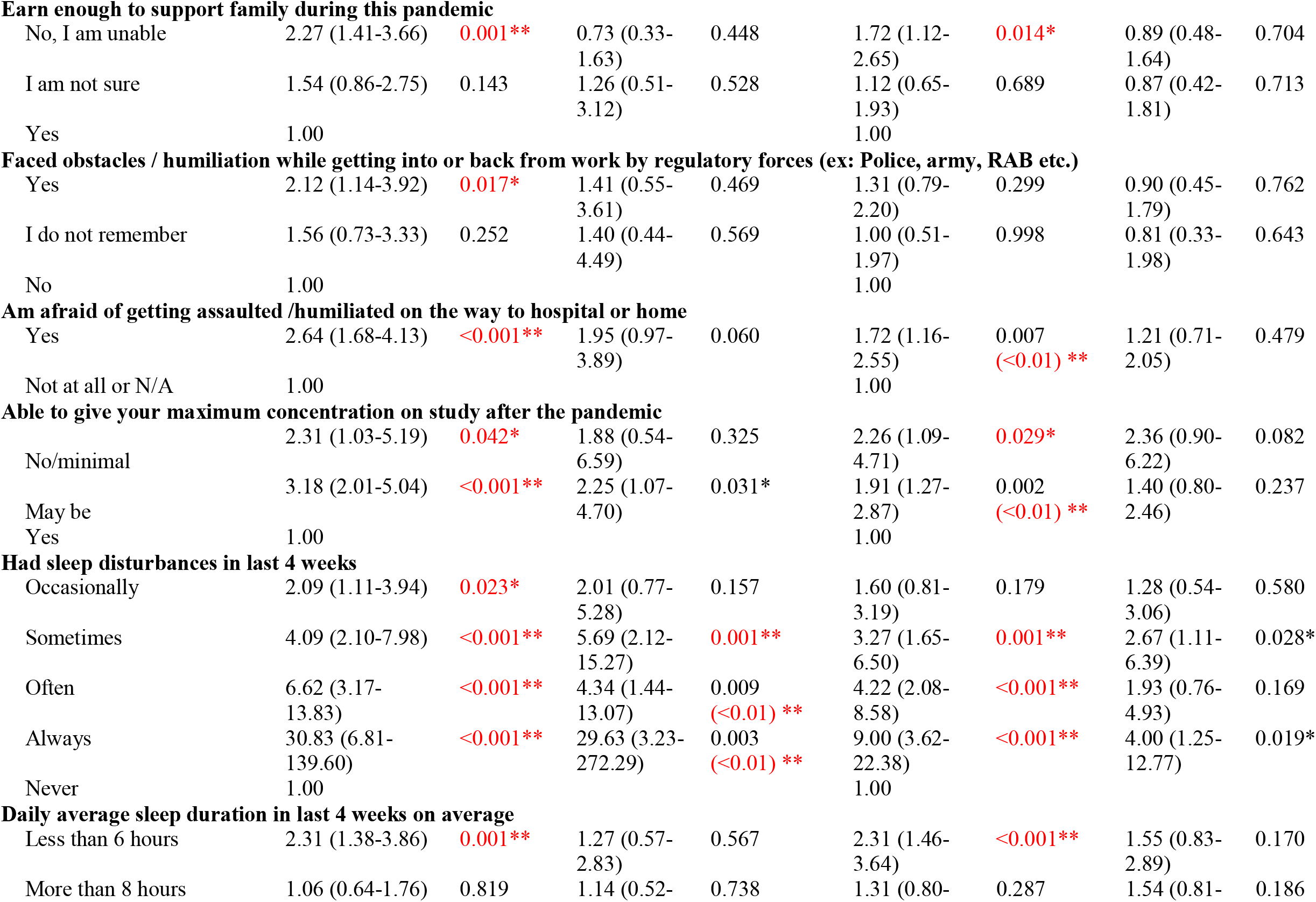
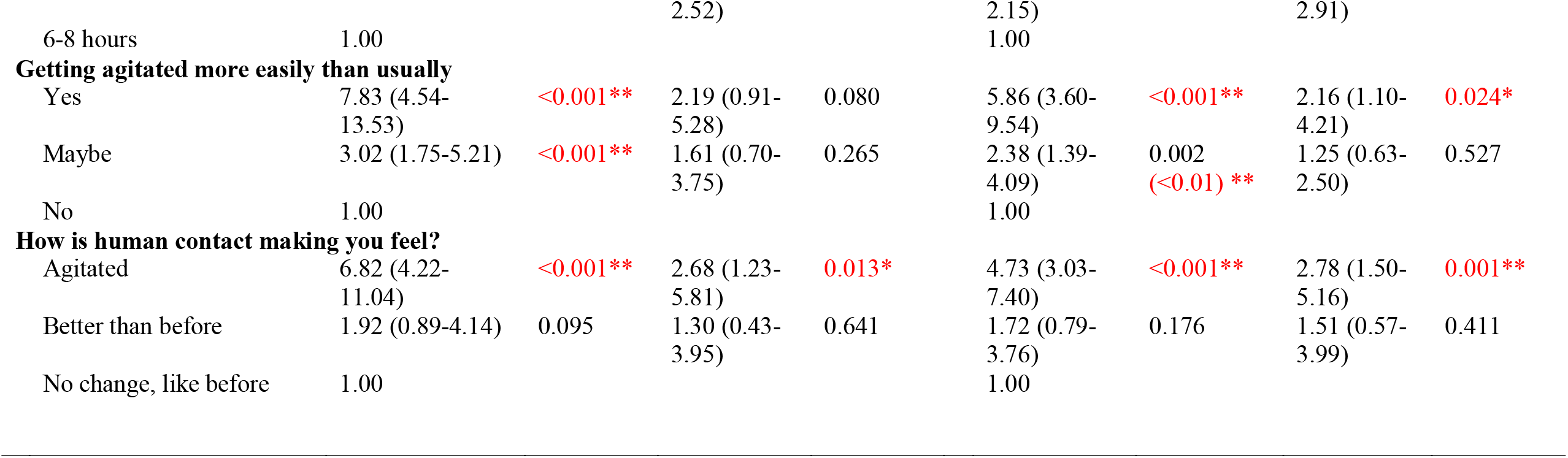
Odds of binary logistic regression of predictive study variables with anxiety and depression among physicians in Bangladesh.

Apart from these, respondents being moderately or severely tensed about being infected by COVID-19,not getting any incentives for patient treatment, feeling that they had not been provided with enough training about COVID-19,not being ready to deal a COVID-19 patient, having news updates from various sources (e.g.TV news, social media, online or offline newspapers etc.) and having sleep disturbances from occasionally to always had significant association with higher odds of either anxiety or depression or both in some cases. (Table 3)

## DISCUSSION

This cross-sectional study investigated the prevalence of anxiety and depressive symptoms in registered Bangladeshi physicians amid the COVID-19 pandemic, and to identify the factors associated with these psychological issues. To the best of our knowledge, no previous studies have taken place among physicians in context of Bangladesh to determine the level of anxiety and depression and their contributing factors related to this pandemic.

Our findings revealed that, in terms of HADS cut-off points, the prevalence of anxiety and depression among registered physicians was 67.72% and 48.54% respectively. Several risk factors were found to be associated with this higher prevalence of depression and anxiety. For instance, among female gender, physicians who had experienced symptoms of COVID-19, not received incentives/only received compliments, reliance on self-funded PPE, inadequate training, lacking perceived self-efficacy while helping COVID positive patients, greater perceived stress of being infected, fear of getting assaulted/humiliated, use of social media, lower income level to support family, feeling more agitated, being agitated while contacting other people, less than 2 hours of leisure activity a day, assuming no positive outcome/impact in life, sleep disturbance, and short sleep duration were found to be positively associated with physicians’ anxiety and depression symptoms in the unadjusted and in the adjusted statistical models.

Worldwide, throughout the pandemic front-line physicians are not only at the risk of physical challenges but also of experiencing significant mental health and psychosocial issues.^13 14^ In line with our findings, an estimated 50.4% and 44.6% healthcare staffs self-reported anxiety and depression respectively in a cross-sectional survey in China.^15^ On the other hand, a significantly lower prevalence of anxiety and depression (14.5% and 8.9% respectively) have been found in a Singapore-based study involving physicians.^16^ The variances between study results could be explained by methodological differences, level of resources available in each of those countries and the subsequent demands on physicians and adoption of different scales and cut-off scores in different surveys, as well as true differences.

One Iranian study utilized the same research instrument (HADS) as our study. In line with our study findings, more than 68% of doctors and nurses had experienced anxiety symptoms, whereas depressive symptoms were reported approximately 52%.^17^ Overall, a recent systematic review and meta-analysis quantified the cumulative prevalence of anxiety and depression experienced by medical staff during COVID-19 by pooling data from 13 studies with a reported 23.2% anxiety and 22.8% depression.^18^ The disproportionately higher prevalence of anxiety and depression (67.72% and 48.54% respectively) among Bangladeshi physicians could be described by the extreme shortage and mal-distribution of health workforce,^19^ coupled with significantly higher rates of infection and death among health professionals, extended working hours, ultimate shortage of personal protective equipment (PPE), lack of adequate training, and social assault/humiliation.

As expected, Bangladeshi female physicians experienced higher psychological distress than their male counterparts during this pandemic. This finding agrees with the established gender gap for more frequent anxious and depressive symptoms among women.^20 21^ In parallel with Wang et al., (2020) study findings, where three-fold higher anxiety disorder was observed among women,^22^ the current study identified females suffering from anxiety symptoms 2.5 times greater than their counterparts. Biological mechanisms and hormonal influences may demonstrate the relationship of higher perceived psychological distress in women ^20^. Although age and associated physical and mental comorbidities are identified as major predisposing factors for anxiety and depression among doctors,^23-25^ surprisingly, we found no statistically significant differences between depression symptoms and anxiety levels and the above-mentioned variables, which warrants future research.

Our study revealed that, less than 6 hours of sleep compared to normal sleep duration (6-8 hours) and any level of sleep disturbances experienced in the past four weeks in comparison to no sleep disturbance, were linked to prevalence of anxiety and depressive symptoms. A survey conducted by Wang et al., (2020) among Chinese pediatric physicians, found independent association between sleep disturbances and depression, however, anxiety reveled statistically non-significant relationship, although physicians with sleep problems reported higher anxiety than their counterparts.^26^ Another study exhibited similar findings that represented Taiwanese local physicians during SARS pandemic.^27^ Moreover, depression, but not anxiety, was found approximately 4 times higher among physicians those spent less than 2 hours on leisure activities than that of 4-6 hours in this current study.

Following COVID-19 outbreak, physicians took initiatives to support the strained health sector and struggled to protect the health of the public,^28^ however, they became the foremost victims of this pandemic concerning the exaggerated psychological pressure of varying factors. For example, direct contact with COVID positive patients, suspected patients hiding medical history, concern about inadequate PPE, extended working hours, infection of colleagues, separation from family, fear of infecting family members, physical fatigue, and medical violence may in turn accelerate their existing stress level.^14 29-31^ Along with these, Lu et al., (2020) emphasized on worrying about being infected, duty in the isolation ward, feeling lonely due to detachment from the loved ones, being frustrated with unsatisfactory results on work, and the fear of an uncontrollable epidemic, leading to psychological distress.^32^ The level of anxiety and depressive symptoms even more prominent among Bangladeshi doctors, contributed by several factors, such as self-funded PPE, absence of incentives, lack of proper training to deal with COVID patients, perceived stress of being infected, and fear of getting assaulted/humiliated while returning home from workplace. Addressing the above-mentioned factors by the policymakers and organizational authorities are paramount to excel the strength of HCWs and support their mental well-being to reach to their highest level of aspirations to serve the humanity.

This study provides novel findings on anxiety and depressive symptoms among Bangladeshi physicians during COVID-19 pandemic, however we cannot overlook the limitations. The cross-sectional nature of the study design could not establish causal relationship between the dependent and independent variables. This study was carried out by conducting a web-based survey, which might generate sampling bias by excluding the physicians who do not have access to internet or inactive in social medias, and thus limit the generalizability of the findings. Besides, self-reported responses on anxiety and depression symptoms only provided subjective data which may greatly differ from objective data, leading to response bias. Finally, although we tried to address major risk factors, several relevant variables, such as residence status (urban or rural), having children, domestic violence, moral dilemma to manage such complex patients and information on physician’s work hours or perceived workloads were not included in the survey.

Despite these limitations, our study has several clear public health implications. Our results suggest vulnerability of Bangladeshi physicians for anxiety and depressive symptoms during the pandemic which should be closely monitored. Previous studies emphasized on alarmingly higher rates of ‘physician burnout’, characterized by emotional exhaustion, depersonalization, and low personal accomplishments,^33^ and alternatively increased risk of suicidal ideation and suicidal attempts in physicians.^34^ Moreover, Montemurro reported suicides in India and Italy during this pandemic, as physicians experienced helplessness, acute psychological stress, and utmost fear of dying.^35^ As physician’s psychological health and patient safety and satisfaction are inextricably linked,^36^ promotion of mental well-being of physician is paramount and there is an urgent call for personal, social, and policy level interventions before it is too late. Given the importance of the risk factors associated with physician’s anxiety and depression symptoms identified in this study, provision of adequate PPE, proper training before deployment in the isolation ward, additional incentives, and on-going monitoring and remote psychological support may aid in reducing physician’s psychological strain.

## CONCLUSION

This study reports a real concern about the prevalence of anxiety and depression symptoms with identification of associated factors among Bangladeshi physicians during the COVID-19 pandemic. Such mental health difficulties are higher than normal scenario. Given the vulnerability of the physicians and other health care staff in this extraordinary condition whilst they are shouldering the overwhelming weight of the epidemic, fighting social stigma and putting their lives at risk to help the affected, health authorities should addressing their psychological needs and formulate effective strategies, SOPs and appropriate interventions to support these frontline fighters at such difficult times.

## Data Availability

Data will be available upon reasonable request.

## Acknowledgement

The authors would like to express their gratitude to Jannatul Ferdous, Wasi Uddin Ahmad, Sakib Hasan, Iftekhar Ahmed Sakib, Lubana Nasreen Tushi, Syed Ramiz Ahnaf, Al Hasnat Turab, Faria Islam Ria, Sajibur Rahman and Md Asifur Rahman for their support in collecting data. Authors would also like to thank all the participants for their spontaneous and voluntary participation in the study. GT is supported by the National Institute for Health Research (NIHR) Applied Research Collaboration South London at King’s College London NHS Foundation Trust, and by the NIHR Asset Global Health Unit award. The views expressed are those of the author(s) and not necessarily those of the NHS, the NIHR or the Department of Health and Social Care. GT also receives support from the National Institute of Mental Health of the National Institutes of Health under award number R01MH100470 (Cobalt study). GT is supported by the UK Medical Research Council in relation the Emilia (MR/S001255/1) and Indigo Partnership (MR/R023697/1) awards.

## Authors contribution

Conceptualization: M.Tasdik Hasan, Syeda Fatema Alam, Md. Abdur Rafi, Vivek Podder, S.M Quamrul Akther, Fatema Ashraf. Data analysis: Afifa Anjum, Sahadat Hossain. Investigation: M. Tasdik Hasan, Syeda Fatema Alam, Md. Abdur Rafi, Vivek Podder, Dewan Tasnia Azad, Rhedeya Nury Nodi. Methodology: M. Tasdik Hasan, Sahadat Hossain. Resources: M. Tasdik Hasan, Farhana Safa, Syeda Fatema Alam, Sahadat Hossain, Supervision: S.M Quamrul Akther, Fatema Ashraf. Writing – original draft: M. Tasdik Hasan, Farhana Safa, Abid Hasan Khan, Afifa Anjum, Sahadat Hossain. Writing – review and editing: M. Tasdik Hasan, Shahadat Hossain, Afifa Anjum, Farhana Safa, Abid Hasan Khan, Kamrun Nahar Koly, Syeda Fatema Alam, Md. Abdur Rafi, Vivek Podder, Tonima Islam Trisa, Rhedeya Nury Nodi, Dewan Tasnia Azad, Fatema Ashraf, S.M Quamrul Akther, Helal Uddin Ahmed, Simon Rosenbaum, Graham Thornicroft. All authors read and approved the final manuscript.

## Patient and Public Involvement statement

Patients and/or the public were not involved in this study.

## Patient consent for publication

Not required.

## Availability of data and materials

Data will be available upon reasonable request.

